# Fatigue Symptoms Influence Effort-Based Decision-Making in Major Depressive Disorder

**DOI:** 10.1101/2025.10.30.25338913

**Authors:** Grace E. Steward, Adam J. Culbreth, Fernando S. Goes, Vikram S. Chib

**Author notes:** Correspondence and requests for materials should be addressed to: Vikram S. Chib, 716 North Broadway, Rm 241, Baltimore, MD 21205, USA.

## Abstract

**Background:** Fatigue is a central symptom of major depressive disorder (MDD). Research in healthy individuals has shown that heightened feelings of fatigue are associated with a reduced willingness to exert effort. However, it remains unclear how fatigue affects effort-based decision-making in individuals with MDD. In this study, we explore how cognitive effort is traded for rewards in MDD and how feelings of fatigue and depressed mood symptoms influence this decision-making process.

**Methods:** Healthy participants (n = 26) and individuals with MDD (n = 18) took part in a forced-choice paradigm, where they decided to perform either a low-effort cognitive effort task for a small reward or a more cognitively effortful task for a larger reward. Participants also completed the Beck Depression Inventory and the Modified Fatigue Impact Scale, which were used in a factor analysis to generate combined scores for depressed mood and fatigue for each participant. These factor scores were then incorporated into hierarchical mixed-effects models of choice data to explain behavioral differences between participants with MDD.

**Results:** Our findings reveal that individuals with MDD exhibited significantly higher preferences for low-effort/low-reward options compared to their healthy counterparts. This difference was linked to increased feelings of fatigue, which raised the perceived cost of cognitive effort. Variations in fatigue questionnaire scores showed stronger associations with effort-based choice behavior than those from questionnaires assessing depressive mood, indicating that fatigue is a key symptom with specific ties to effort in MDD.

**Conclusions:** These findings illuminate how fatigue might lead to decreases in goal-directed behavior in MDD, thereby deepening our understanding of diminished motivation in MDD and suggesting potential pathways for more effective treatment.

## INTRODUCTION

MDD profoundly affects various aspects of individuals’ lives, including motivation, decision-making, and willingness to exert effort. Diagnostic criteria of MDD include fatigue as a core symptom^1^, which can impact these behaviors, disrupting the ability to engage in tasks that require cognitive effort. While there has been a great deal of work to examine how MDD influences decisions to exert effort, and fatigue is a well-established symptom, there is a limited understanding of how MDD-induced fatigue impacts decision-making and motivated performance. In this study, we examine how individuals with MDD trade cognitive effort for rewards and how fatigue and depressive symptoms shape this decision-making process.

Effort-based decision-making has been proposed as a behavioral approach for understanding motivation. In this framework, the willingness to exert effort is influenced by the subjective value of the rewards associated with a particular outcome, as well as the trade-off between the reward and the effort required to obtain it. Effort-based decision-making studies have been applied in the context of physical^2–8^ and cognitive^8–11^ effort in healthy individuals, revealing that the willingness to undertake effort increases as the levels of prospective effort-contingent reward rise, and that the subjective value of effort increases through repeated fatigue-induced exertion.

Recently, several studies have explored cognitive and physical effort-based decision-making in individuals with MDD. While these works have reported some mixed results^12–15^, generally, it has been found that individuals with MDD are less willing to exert cognitive^16–20^ and physical^15,19–25^ effort for monetary rewards compared to healthy individuals. Although studies in effort-based decision-making have shown that feelings of fatigue have a significant effect on effort-based decision-making^2,6,7,9,26,27^, and fatigue is a hallmark symptom of depression, there has been limited investigation of how symptoms of fatigue may influence effort-based decision-making in individuals with MDD.

In this study, we investigate differences in cognitive effort-based decision-making between individuals with MDD and healthy controls while evaluating whether fatigue or depressed mood provide a better explanation for these behavioral differences. We hypothesize that individuals with MDD would be less willing to exert the same cognitive effort for monetary rewards when compared to healthy controls. Additionally, we expect that fatigue symptoms in MDD will significantly influence these decision-making differences and offer greater insight into effort-based decision making than depression symptoms alone. These hypotheses build on prior research examining decision-making in individuals with MDD, along with studies of fatigue in healthy individuals, to provide an account of how fatigue might influence choice in individuals with depression.

## METHODS AND MATERIALS

### Participants

Participants for this Johns Hopkins School of Medicine Institutional Review Board-approved study were recruited from the Johns Hopkins University community via online postings. We collected data from 21 participants meeting DSM-5 criteria for major depressive disorder (with no history of psychosis), and 29 healthy participants from the same community with no self-reported history of neurological or psychiatric disorders. Participants were excluded if they met the criteria for substance dependence or abuse evaluated under the Mini-International Neuropsychiatric Interview^28^. Medication history was collected from the MDD cohort (Table S1). No participants in the control cohort reported having ever taken a psychotropic medication. Informed consent was obtained before the commencement of the study.

After collection, participant data were excluded if their choices indicated aversion to reward during the experiment. This was determined using a logistic regression of choice data. Participants with a negative weight for reward were excluded. Three participants were removed from each group, leaving a total of 18 participants with MDD and 26 healthy controls.

There were no significant differences in age, sex, race, ethnicity, or income between cohorts (**Error! Reference source not found.**). However, a marginal trend level difference in years of education was found between groups (t(42) = −1.867, P = 0.069, with those in the MDD group having nominally less years of education). As a result, we conducted a sensitivity analysis to examine the differences between groups, focusing on the variations in education level. We did not find a significant effect of education level on choice behavior between the two groups (Table S3; t(1038) = 0.474, P = 0.635).

#### Diagnostic and Symptom Assessment

Diagnoses in MDD cases were performed using the Diagnostic Interview for Genetics Studies (DIGS) version 4.0^29^ based on DSM-4 criteria. DIGS assessments were not performed on healthy control participants. To assess symptoms of depression and fatigue, all participants completed the Beck Depression Inventory (BDI)^30^ and the Modified Fatigue Impact Scale (MFIS)^31^.

### Cognitive Effort Task

Before the study began, participants were informed that they would receive a $15 show-up fee, regardless of their performance during the task, but that they would also earn additional money based on their choices and performance. Our paradigm design was influenced by a previously designed cognitive effort task from Westbrook *et al*.^10^However, we made adjustments in the choice phase of the paradigm so that the range of reward and effort was not explicitly linked in a progressive ratio task. In this way, effort levels and rewards were orthogonalized in our experimental paradigm, allowing us to analyze the effects of reward and effort separately^9^. We began our experiment by familiarizing participants with the cognitive effort needed to complete a level of the *n*-back task (Figure 1A). Each *n*-back task consisted of 40 sequentially presented letters. Within this sequence, there were 10 target letters that a participant had to identify as the same letter *n* letters prior. Participants had approximately 2 seconds to identify whether the current letter on the screen was one of these targets. Once a participant entered their choice, the next letter would be presented. Participants had to correctly identify 50% of these targets to succeed in the task. Feedback was presented at the end of each sequence. If participants correctly identified 50% of the targets, they received a “Success!” message on the screen. If they failed to achieve 50% success, they received a message of “Please try harder!” and continued to the next n-back level. To avoid anchoring effects of subsequent difficulty ratings to numeric effort levels, participants associated each of the four *n*-back task levels, where *n* ranges from 1 to 4, with a distinct color. Participants experienced a single effort level three times successively. Each effort level was presented in random order without replacement.

**Figure 1.**
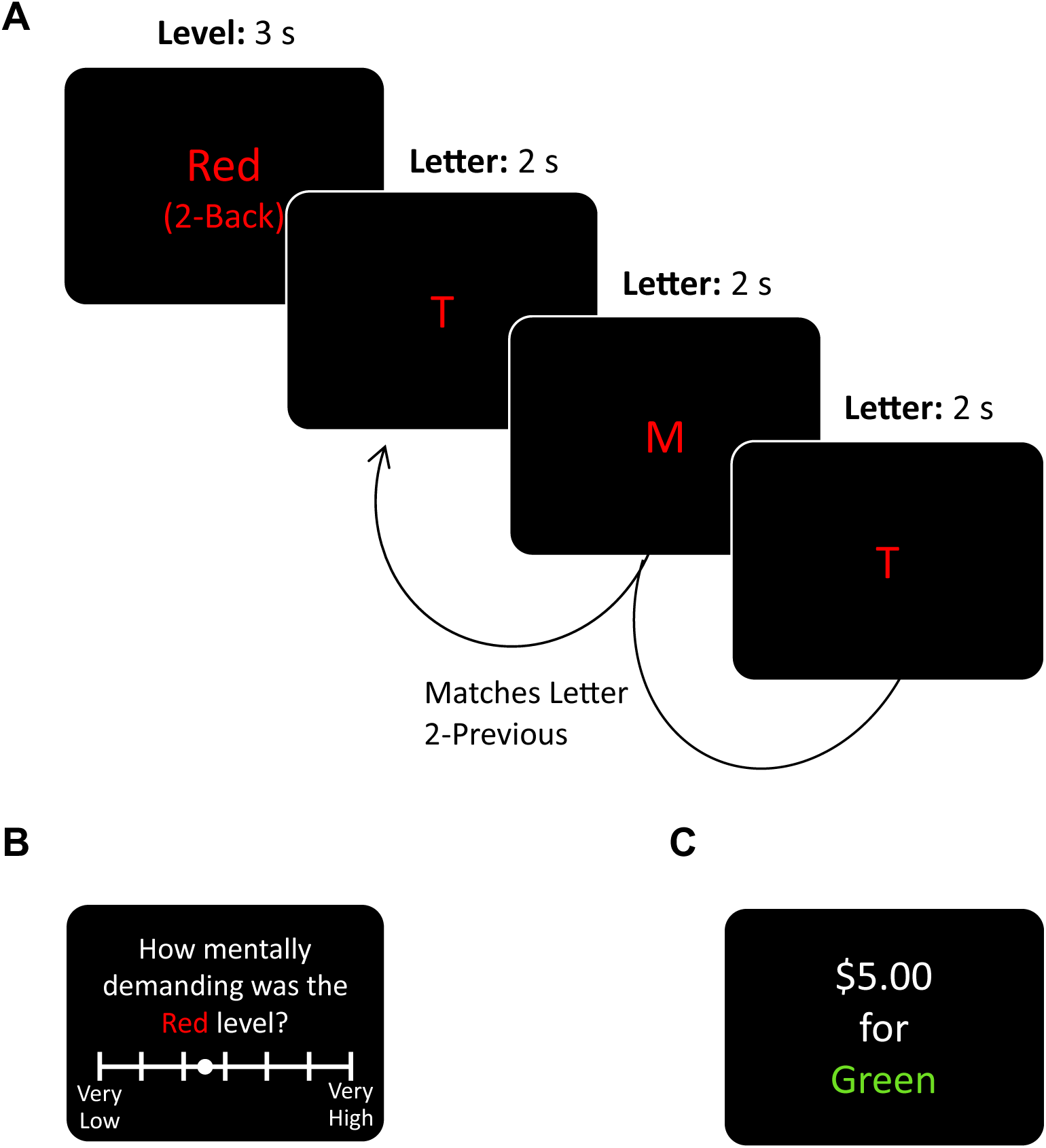
Experimental Design. **A.** Participants first associated different effort levels of the *n*-back task with distinct colors. During the n-back cognitive effort task, participants identified when a letter presented on the screen matched the one presented *n* letters previously. **B.** Following this association phase, participants rated the difficulty of each color level on a continuous scale from ‘Very Low’ to ‘Very High’ mental demand. **C.** During choice trials, participants chose between a default effort/reward (1 back for $1) option and an option with varying levels of effort for different reward amounts.

Following this association phase, participants underwent an assessment phase to ensure that they had developed salient associations between levels of cognitive effort and colors. They rated the mental challenge of each *n*-back level on a continuous scale from “Very Low” to “Very High” (Figure 1B).

Finally, participants underwent a choice phase in which they could opt for a default option, to complete a 1-back task for $1, or engage in a variable level of cognitive effort for a variable monetary reward displayed on the screen (Figure 1C). This phase consisted of 24 unique offers that orthogonally spanned three effort levels and monetary rewards ranging from $1 to $8, in increments of $1 (Table S4). To ensure this choice paradigm was incentive compatible, at the end of the experiment, two offers from the choice phase were randomly selected and realized. The participant then executed the chosen option, exerting the associated effort and earning the reward. Participants needed to succeed in the effort task to receive the reward, and they were given three opportunities to succeed.

### Data Analysis

#### Task Performance

Task accuracy was calculated as the ratio of how many targets and nontargets were properly identified out of 40 total letters in each *n*-back trial. To evaluate differences in task accuracy between MDD and control groups, we constructed a linear mixed-effects model that related task accuracy to *n*-back level with group as a categorical fixed effect and participant as a random effect to account for inter-individual differences:

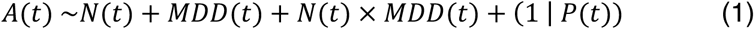

Parameters were estimated using the *fitlme* function of MATLAB 2023a^32^, with relative coding of dummy variables and the maximum likelihood fit method. This model included task accuracy *A(t)* and *n*-back level *N(t)* as continuous variables, while *MDD (t)* represents a categorical variable of the diagnostic group (MDD/Control), and *P(t)* is a categorical variable for each participant for a particular trial *t*.

#### Task Difficulty Ratings

To analyze differences in task difficulty ratings between cohorts, we constructed a similar confirmatory linear mixed-effects model:

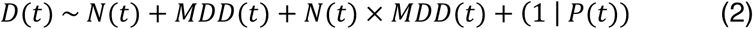

This model included task difficulty ratings *D(t)* and *n*-back level *N(t)* as continuous variables. Mirroring the task-performance model, *MDD(t)* represents a categorical variable of the diagnostic group and *P(t)* are categorical identifiers for a participant of a given trial *t*. This model was estimated using the same model estimation procedure as the task-performance model.

#### Analysis of Between-Group Choice Differences

To test if there was a difference in choice behavior between the MDD and control cohorts, we conducted a confirmatory analysis of the effect of group on decisions made during the choice phase. We modeled choice data using a hierarchical generalized linear mixed-effects model to capture all available data from the choice trials, while accounting for individual-participant differences. We assumed that choice would be affected by the effort and reward of the presented option and that there would be an effect of cohort on choice, all of which were modeled as fixed effects. We expected random biases attributable to each participant, modeled as a random intercept:

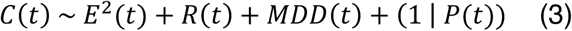

where *C(t)* is the choice being made on a given trial *t* (0 = default option, 1 = presented option), *E(t)* is the effort of the presented option encoded as the *n* in the *n*-back being offered. The square of the effort was used in accordance with previous models of the *n*-back task^9,33,34^, *R(t)* is the reward corresponding to the successful completion of the effort level, *MDD(t)* is a categorical variable of group, and *P(t)* is a categorical participant identifier. Effort and reward were linearly standardized on the interval of [0,1] to preserve the significance of sign. Models were estimated with the *fitglme* function of MATLAB 2023a^32^ using the Laplace approximation of maximum likelihood^35,36^ and an assumption of a binomial distribution of the binary response variable of choice.

We used bootstrapping to create confidence intervals for parameter estimates because it was unlikely that the underlying distribution of the beta weights associated with the cohort was strictly normal, given the sample’s makeup of individuals with varied symptom severity. We created 2000 pseudo datasets by sampling each participant with replacement to generate pseudo participants, with 24 choices for each subject. Parameters were estimated from these pseudo datasets and used to create a distribution of the sample parameters.

#### Factor Scores of Fatigue and Mood Symptoms

To examine which aspects of choice could be linked to fatigue and mood symptoms, we performed an exploratory factor analysis (EFA) in R v4.2.2^37–39^. Because the BDI and MFIS scales included two target dimensions, we assumed a two-factor structure (BDI: depression severity; MFIS: fatigue severity). We chose EFA to identify these latent variables because EFA assumes an underlying structure and causality with latent factors, unlike other techniques like principal component analysis (PCA), which are more suited to dimension reduction and visualization^40^. We conducted three factor analyses using different item-retention criteria to strike a balance between data retention, reduction of multicollinearity and cross-factor loading, item suitability, and stability of the factor structure. The analysis described here was sued for hypothesis testing, and additional models are documented in the Supplemental Materials.

We used symptom scale data from both groups and first tested our data for instances of multicollinearity, which can cause issues of singularity in estimation^41^. This was achieved by calculating the Pearson correlations between each item given the responses of all participants (Table S10). We removed items with cross-correlations higher that 0.8 with another item (BDI Items 4 and 15, and MFIS Items 15,19, 21). Over-correlated items were removed such that all item correlations fell below a 0.8 correlation threshold^41^.

We then ran an initial factor analysis and removed items with cross-factor loadings that had less than a 0.1 loading difference between the two factors^42^. We repeated the process until no items cross-loaded onto the two factors^40^, resulting in removal of items (BDI 5, 7, 8, 14, 17 and MFIS 18). This was done to improve the interpretability of factors and factor scores.

We verified that our remaining data were sufficient for factor analysis and conducted Bartlett’s test of sphericity and Kaiser‒Meyer‒Olkin (KMO) test measure of sampling adequacy of our data using the *cortest.bartlett()* and *KMO()* functions, respectively.

Given that our data met the criteria for adequacy (Bartlett’s test of sphericity, χ^2^(465) = 1562.6, P = 1.24E-118, KMO, Overall MSA = 0.75), we conducted a factor analysis using the *factanal* function within the *stats* v4.2.2 base library package. We assumed two latent factors based on the targeted dimensions of the BDI and MFIS. Parallel analysis and the resulting scree plot of eigenvalues suggested that one factor may have been sufficient but given that the goal of this factor analysis was to generate factor scores associated with fatigue and depressed mood symptoms and that scales were designed to test two distinct domains, we assumed a 2-factor structure.

Factor analysis was completed using a ‘*promax*’ oblique rotation, as fatigue and mood symptoms are highly correlated. Bartlett factor scores were generated from this analysis for each participant and were used in the analysis of within-diagnostic group choice (Table S15)^43^.

#### Analysis of Within Diagnostic Group Choice

Given the difference in behavior between the two cohorts, and to assess the impact of various levels of clinically significant depression on behavior, we modeled the MDD cohort without the control cohort. Using data from the control cohort, including depression and fatigue assessments, would cause issues related to a lack of variability in their BDI and MFIS assessments. To determine the best explanation of the behavior in the MDD cohort, we began with a null model in which participant choice was based on the level of effort and reward offered.

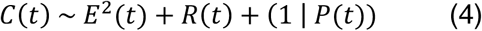

Using a forward method, we estimated more complex models that incorporated a participant’s individual factor score for both fatigue and depressed mood. Using the model with the lowest AIC from the group, we conducted a likelihood ratio test between the best-fit model and the null model to determine if the fit was significantly better than that of the null model.

Models were estimated with the *fitglme* function of MATLAB 2023a^32^ using the Laplace approximation of maximum likelihood^35,36^ and an assumption of a binomial distribution of the binary response variable of choice. We then followed the same bootstrapping procedure used in the evaluation of differences in choice due to diagnostic cohort.

## RESULTS

### Differences in Choice Preferences Between Individuals with MDD and Healthy Participants

To evaluate differences in effort perception or task performance between the MDD and control groups, we analyzed both cognitive exertion performance and effort perception across the levels of the *n-*back task. We did not find significant differences in task accuracy (Figure 2A; linear mixed effects model, t(173) = −0.257, P = 0.798) or the perception of task difficulty (Figure 2B; linear mixed effects model, t(173) = 0.984, P = 0.327) between groups. These results suggest that performance and perception of cognitive effort were matched between groups, allowing us to make inferences about differences in choice between groups that are not the byproduct of differences in perception or performance.

**Figure 2.**
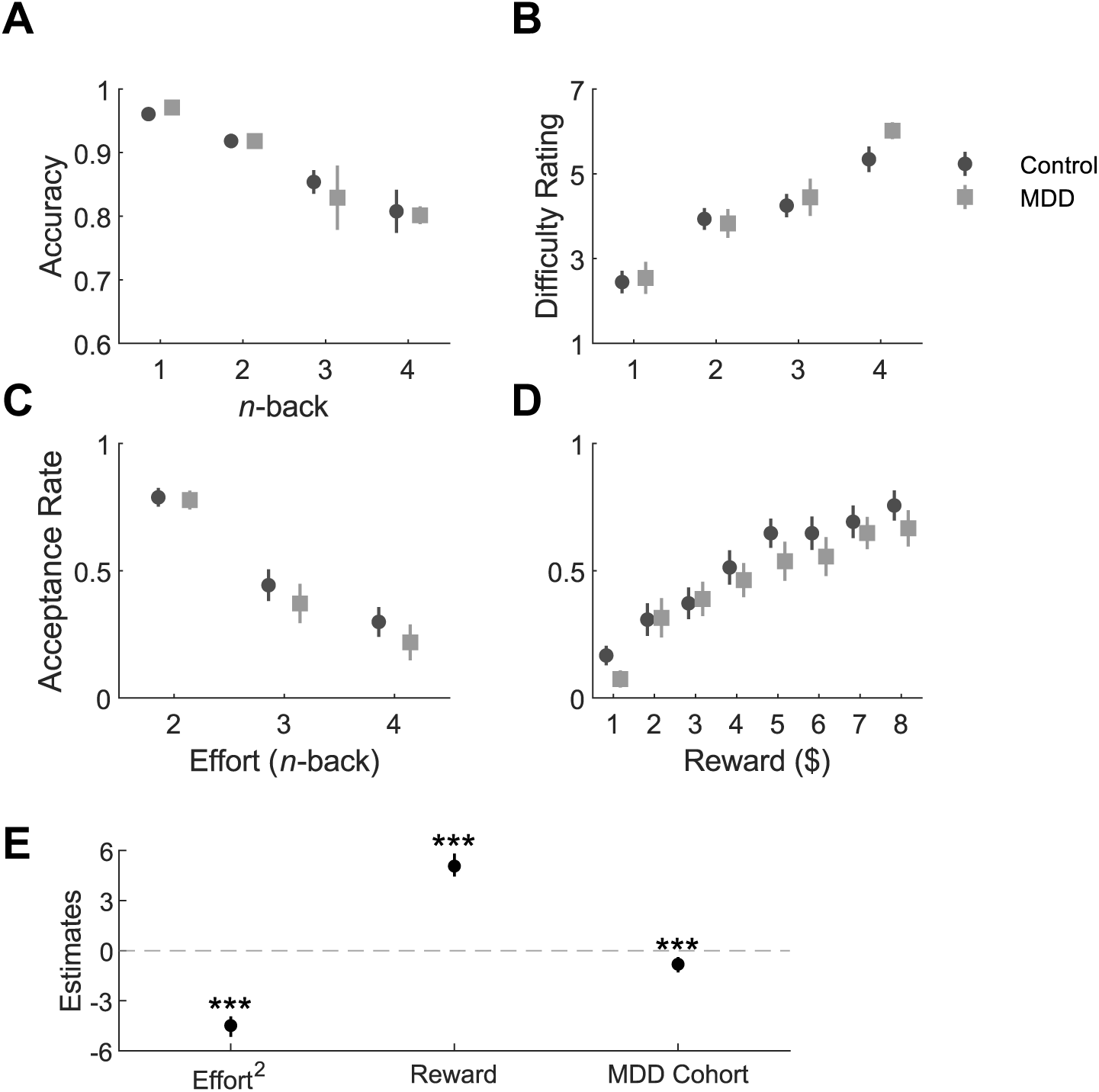
Behavioral Data. **A.** Working memory task accuracy as a function of *n*-back level for the MDD and control groups. **B.** Participants’ ratings of working memory task difficulty as a function of the n-back level, during trials, polling mental demand. Probability of accepting the nondefault effort/reward option as a function of cognitive effort level **(C)** and reward **(D).** Error bars represent standard error. **E.** A confirmatory mixed-effects model, with bootstrapped parameter estimates displayed, shows a significant bias in the MDD cohort towards the default option compared to their healthy counterparts. *** represents that 99.9% confidence intervals of parameter estimates did not include 0.

Participants in the MDD and control groups’ likelihood to accept the nondefault effort/reward option decreased with increasing effort levels (Figure 2C) and increased with increasing reward (Figure 2D). We found that individuals in the MDD cohort had a greater propensity to select the low-effort/low-reward default option than controls (Figure 2E; 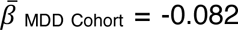, 95% CI = [−1.303, −0.393]).

Evaluating the influence of effort performance and perception on choice, we found that *n-*back level was a better predictor of choice than task accuracy (n-back level AIC = 856; task accuracy AIC = 1169), and that task accuracy did not improve the model in the presence of the *n*-back level variable (Log-likelihood Ratio Test, χ2(1) = 0.65, P = 0.429). These results suggest that any difference in choice behavior between the control and MDD participants is more related to the effort-based decision-making process itself than to differences in the exertion of cognitive effort.

### Separating Fatigue and Mood Symptoms

To evaluate how depression and fatigue symptoms related to choice, we collected responses from the BDI and MFIS. The total scores from both the BDI (t(42) = −9.319, P = 8.832 E −12) and MFIS (t(42) = −7.255, P = 6.284E −12) were significantly different between groups, indicating that MDD participants exhibited higher levels of depression and fatigue than control participants. Within the MDD cohort, we did not find a significant effect of total BDI or total MFIS score on either cognitive task accuracy (BDI: t(69) = 0.717, P = 0.476; MFIS: t(69) = −0.334, P = 0.739) or subjective difficulty rating (BDI: t(69) = −0.024, P = 0.981; MFIS: t(69) = 0.353, P = 0.725). As expected, total BDI and MFIS scores were significantly correlated (ρ = 0.845, P = 1.711 E −13). Thus, assessing whether fatigue reasonably explained choice behavior in MDD in the context of other symptoms, such as anhedonia and amotivation required a more complex analysis.

To separate fatigue and mood symptoms in MDD, we conducted an exploratory factor analysis using the BDI and MFIS survey data. This analysis included two latent factors, one we termed “fatigue”, which was associated with cognitive and physical fatigue symptoms, and the other we termed “depressed mood”, which was most associated with feelings of sadness, lowered social interest, and discouragement (Figure 3A). Factors were allowed to be correlated, with an estimated correlation of 0.76. Both factors accounted for 52.3% of the total variance of the BDI and MFIS scales, while the fatigue and depressed mood factors accounted for 27.6% and 26.3% of the variance, respectively. Two factors sufficiently described the data (χ2(404) = 622.02, P = 1.67E-11). The fatigue and depressed mood factors had reliability values of α = 0.97 and α = 0.93, respectively. Using the resulting factor structure (Figure 3B), we calculated Bartlett Factor Scores for each participant based on their individual survey responses. These factor scores showed that fatigue and depressed mood symptoms are separable and can be used to evaluate choice independently.

**Figure 3.**
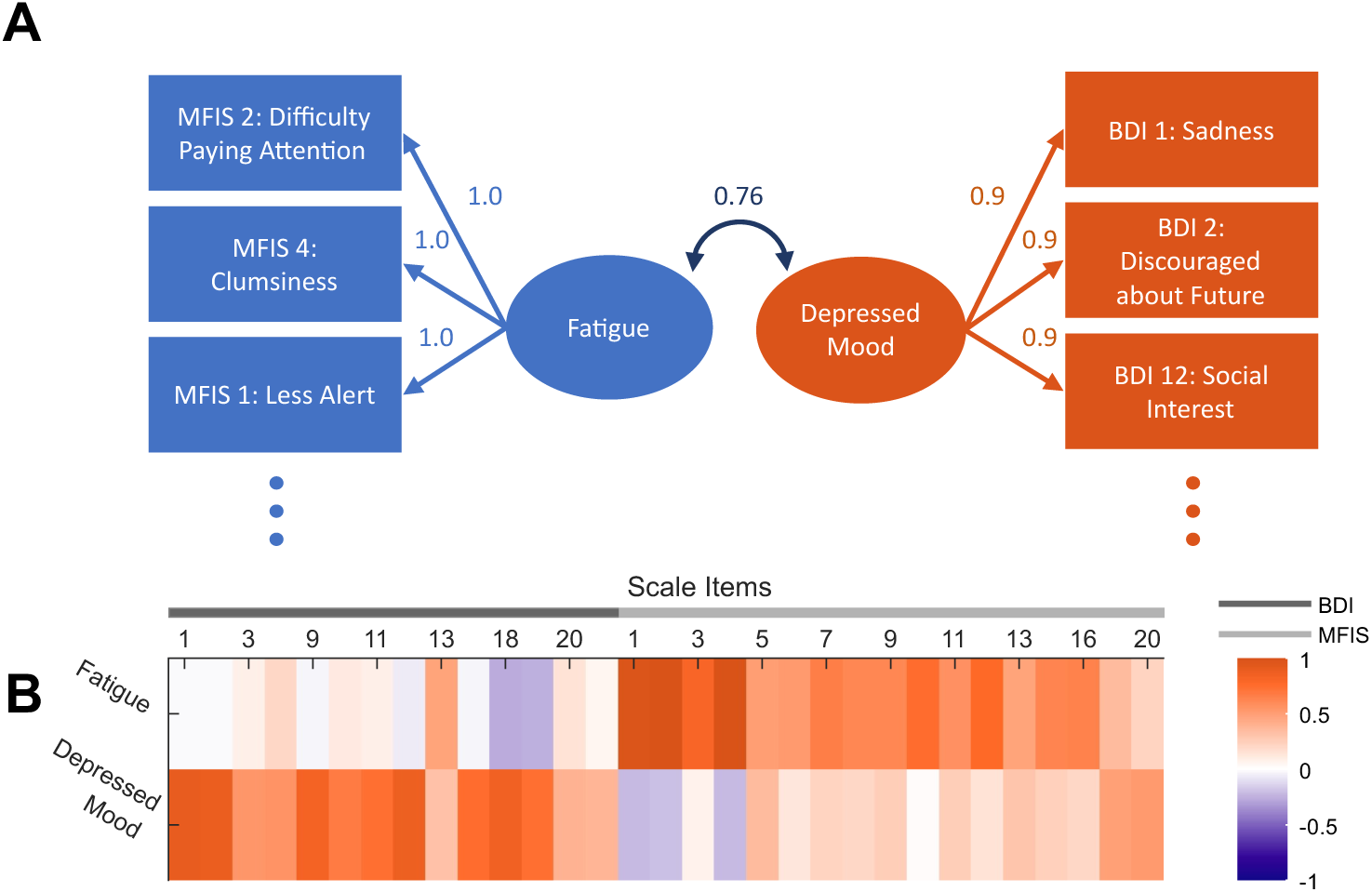
Factor Analysis of Self-Report Fatigue and Depressed Mood Surveys. **A**. Factor loadings of the highest weighted items for each of the two factors. **B**. Factor loadings, with red indicating positive loadings and blue indicating negative loadings, derived from an exploratory factor analysis of responses to the Modified Fatigue Impact Scale (MFIS) and the Beck Depression Inventory (BDI). Two latent factors were estimated, allowing for correlation between them, resulting in distinct factors with item loadings reflecting fatigue and depressed mood.

### Fatigue as a Predictor of Choice within Major Depressive Disorder

To investigate how fatigue and other depressive symptoms might influence differences in choice behavior between the MDD group and controls, we compared model criteria statistics across several models that used either depressed mood or fatigue factor scores as predictors of choice (Figure 4A). Our analysis revealed that the best-fitting model showed a significant interaction between a participant’s fatigue factor score and their subjective cost of effort (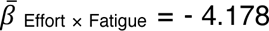, 95% CI = [−7.191, −1.480], Figure 4B). Higher fatigue factor scores were associated with increased perceived cost of prospective effort. This model explained the data better than a null model that considered only effort and reward levels (χ^2^(2) = 7.814, P = 0.020). The winning model outperformed models that included mood symptoms as a factor in choice or models where reward value was influenced by either symptom score (Figure 4A, Table S16).

**Figure 4.**
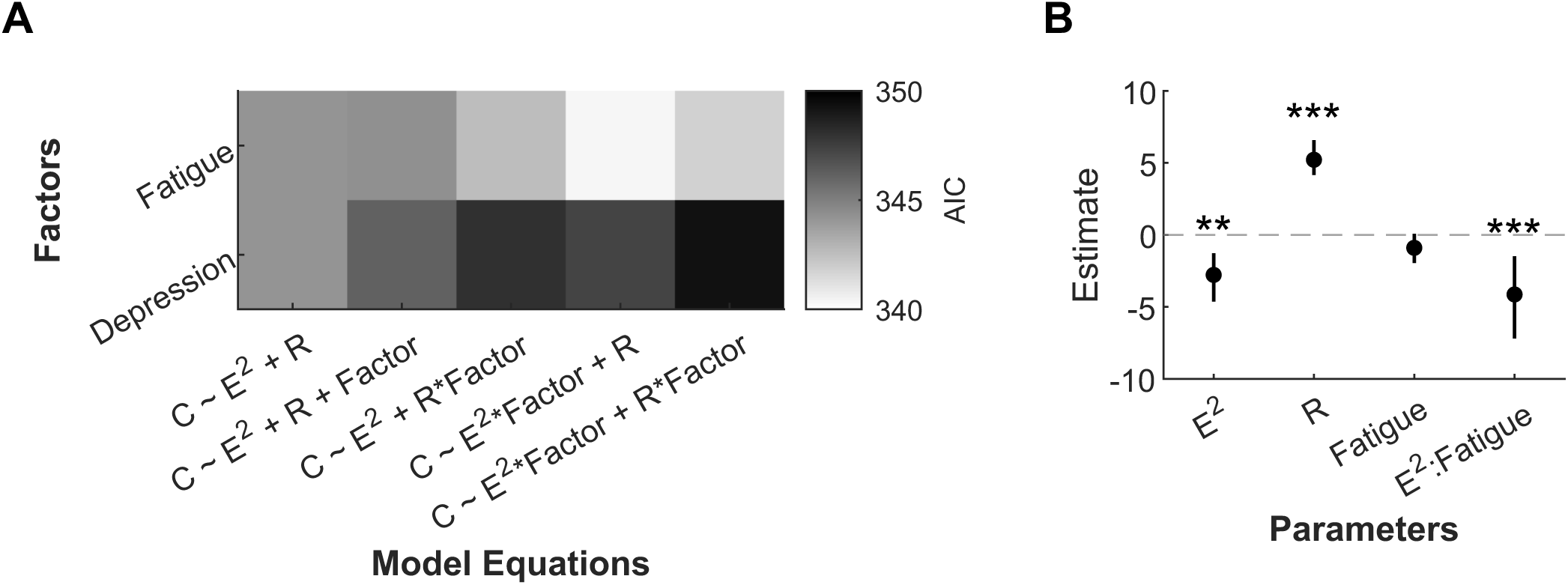
Fatigue best describes choice preferences in MDD. **A**. Participants were assigned individual Bartlett factor scores for Fatigue and Depressed Mood based on their survey responses. These factor severity scores were then used to predict choice behavior. The best-fitting model showed that choice was influenced by fatigue scores, which affected the perception of effort in individuals with MDD. This model performed better than models where either factor impacted the perception of reward. C = Choice, E = Effort, R = Reward. **B**. The best-fitting model indicated that increased fatigue led to higher effort costs. Error bars represent 95% confidence intervals of bootstrapped parameter distributions. ** 99% CI does not include 0; *** 99.9% CI does not include 0.

## DISCUSSION

We examined differences in effort-based decision-making between individuals with MDD and control participants, and assessed how symptoms may influence effort-based decisions. We found that individuals with MDD showed a bias toward choosing the low-reward/low-effort option compared to healthy individuals. Using exploratory factor analysis, we derived symptom factor scores for fatigue and depression symptoms. These scores helped explain differences in effort-based decision-making within the MDD group. Our analysis revealed that a reduced willingness to exert effort in MDD is best explained by fatigue symptoms increasing the perceived cost of prospective effort. Importantly, we did not find evidence that fatigue or depressed mood symptoms lowered reward value in those with MDD. These findings highlight the potential importance of understanding how fatigue impacts motivation in psychiatric and neurological disorders.

Our results align with previous studies that have shown that individuals with MDD have a reduced willingness to exert effort for reward^17,19,20,24,44^. Our paradigm orthogonalized prospective effort and reward, allowing us to isolate their effects on decision-making. Some previous evaluations of cognitive effort costs either used qualitative effort tasks or provided two or fewer effort levels for comparison^17,24,45^. Other studies used performance metrics to understand why those in depressive episodes would exert less effort for the same reward^19^. Those studies may have been less sensitive to differences in effort-based decision-making between individuals, leading to some of the null findings regarding differences in effort valuation between individuals with MDD and control participants^12–14^. The design of our study not only revealed between-group differences but also enabled us to assess how effort cost varies with depressive symptoms.

Previously, work examining effort-cost decision-making in MDD has focused on decreased sensitivity to reward as a central explanation of choice differences between those with depression and their control counterparts^17,20,24^. While the effect of reduced sensitivity to anticipatory and consummatory reward in MDD is well documented^46–49^, we did not find an effect of increased MDD symptom severity on reward value in this study. Instead, in the context of our study, we found that increased effort cost was a significant component of the behavioral difference between groups. These findings are consistent with recent studies that have more fully sampled the space of prospective reward and effort value^18,19^, and support the need for further studies that incorporate different factors that may contribute to effort cost, such as effort perception and task difficulty.

Our study suggests that fatigue plays a significant role in the decision-making of individuals with MDD. Fatigue is a central symptom of MDD^1^, and it has been postulated that it may contribute to other documented symptoms, such as cognitive deficits or inability to disengage from negative stimuli^50^. However, understanding how fatigue-related feelings in MDD contribute to differences in behavior remains understudied. Here, we present evidence that differences in effort-based decision-making in MDD are exacerbated by increased fatigue, rather than mood symptoms alone.

While our study shows a link between fatigue symptoms and changes in decision-making, more research is needed to understand how fatigue impacts this process. Since fatigue is common in many neurological and psychiatric conditions, it is important to determine whether its effects in MDD differ from those in other disorders. In the future, it will be valuable to explore whether the same disruptions that cause fatigue also contribute to other symptoms in MDD, or whether fatigue itself mediates these symptoms. Understanding these dynamics could provide insight into the recently observed transdiagnostic differences in effort-based decision-making among bipolar disorder, schizophrenia, and MDD^12,13,44,51^.

Our study shows that cognitive effort-based decision-making is disrupted in MDD, indicating that effort cost, modulated by fatigue symptoms, significantly affects choice behavior in people with MDD. We distinguish fatigue symptoms from other MDD symptoms and examine their separate roles within the disorder. These findings start to elucidate how fatigue may contribute to disrupted behaviors in psychiatric disorders, thereby enhancing our understanding of depressive conditions and potentially guiding treatment approaches.

## Data Availability

All data produced in the present study are available upon reasonable request to the authors.

## ACKNOWLEDGEMENTS

This work was supported by the National Institutes of Mental Health under Award Number R01MH119086 to V.S.C. and by the Johns Hopkins University Discovery Award to V.S.C. and F.S.G.

## AUTHOR CONTRIBUTION STATEMENT

**G.E.S:** Conceptualization, Methodology, Software, Formal analysis, Investigation, Data Curation, Writing – Original Draft, Writing – Review & Editing, Visualization **A.J.C.:** Writing – Review & Editing, Supervision **F.S.G:** Conceptualization, Methodology, Writing – Review & Editing, Supervision, Funding acquisition **V.S.C:** Conceptualization, Methodology, Writing – Review & Editing, Supervision, Funding acquisition

## DATA & CODE AVAILABILITY

The data generated for this experiment and the code used for its collection and analysis are available on the Open Science Framework at https://osf.io/nfmp5/.

---

**Table 1.**
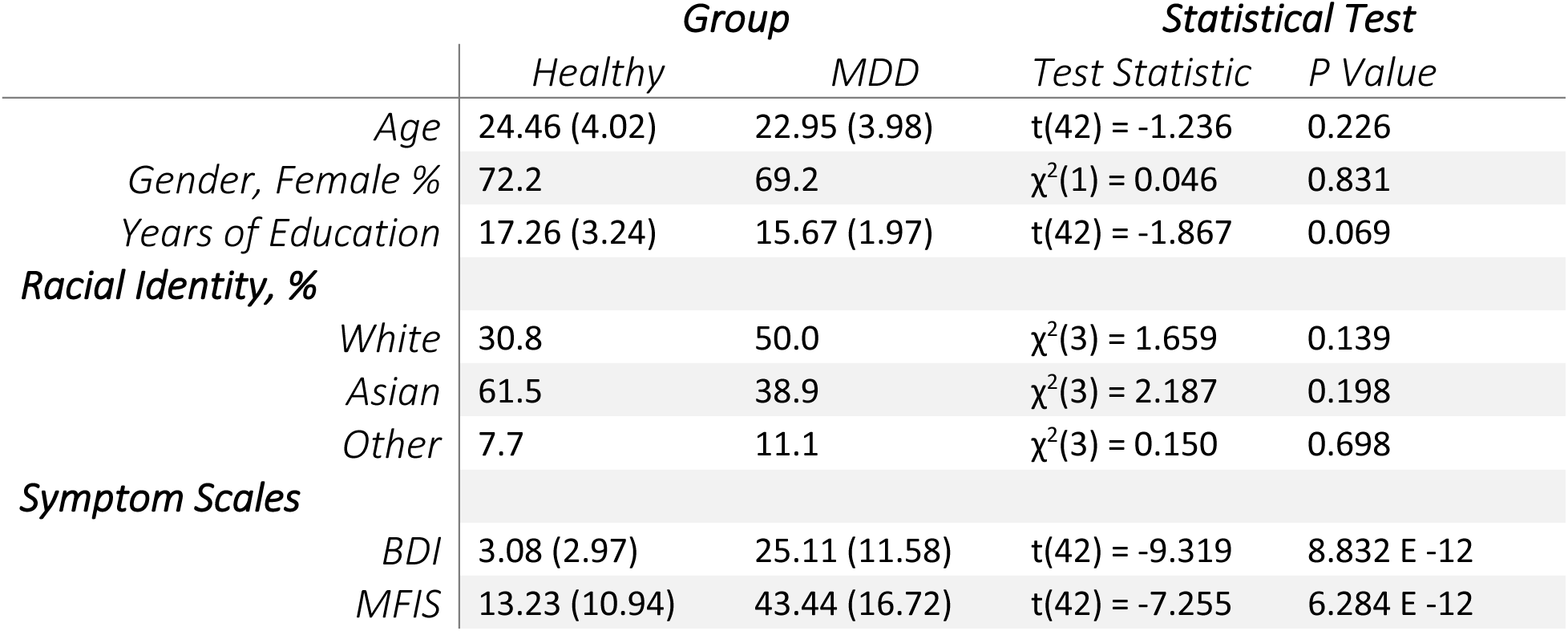
Demographic Summary.

|  | *Group* |  | *Statistical Test* |  |
| --- | --- | --- | --- | --- |
|  | *Healthy* | *MDD* | *Test Statistic* | *P Value* |
| *Age* | 24.46 (4.02) | 22.95 (3.98) | $t(42) = -1.236$ | 0.226 |
| *Gender, Female %* | 72.2 | 69.2 | $\chi^2(1) = 0.046$ | 0.831 |
| *Years of Education* | 17.26 (3.24) | 15.67 (1.97) | $t(42) = -1.867$ | 0.069 |
| *Racial Identity, %* |  |  |  |  |
| *White* | 30.8 | 50.0 | $\chi^2(3) = 1.659$ | 0.139 |
| *Asian* | 61.5 | 38.9 | $\chi^2(3) = 2.187$ | 0.198 |
| *Other* | 7.7 | 11.1 | $\chi^2(3) = 0.150$ | 0.698 |
| *Symptom Scales* |  |  |  |  |
| *BDI* | 3.08 (2.97) | 25.11 (11.58) | $t(42) = -9.319$ | 8.832 E -12 |
| *MFIS* | 13.23 (10.94) | 43.44 (16.72) | $t(42) = -7.255$ | 6.284 E -12 |

## REFERENCES

1. American Psychiatric Association. Diagnostic and Statistical Manual of Mental Disorders. (American Psychiatric Association, 2013). doi:10.1176/appi.books.9780890425596.

2. Hogan, P. S., Chen, S. X., Teh, W. W. & Chib, V. S. Neural mechanisms underlying the effects of physical fatigue on effort-based choice. Nat Commun 11, 4026 (2020).

3. Hogan, P. S., Galaro, J. K. & Chib, V. S. Roles of Ventromedial Prefrontal Cortex and Anterior Cingulate in Subjective Valuation of Prospective Effort. Cerebral Cortex 29, 4277–4290 (2019).

4. Dryzer, M. & Chib, V. S. Neural mechanisms underlying the effects of cognitive fatigue on physical effort-based choice. Preprint at 10.1101/2024.12.06.627274 (2024).

5. Arulpragasam, A. R., Cooper, J. A., Nuutinen, M. R. & Treadway, M. T. Corticoinsular circuits encode subjective value expectation and violation for effortful goal-directed behavior. Proc Natl Acad Sci USA 115, E5233–E5242 (2018).

6. Müller, T., Klein-Flügge, M. C., Manohar, S. G., Husain, M. & Apps, M. A. J. Neural and computational mechanisms of momentary fatigue and persistence in effort-based choice. Nat Commun 12, 4593 (2021).

7. Meyniel, F., Sergent, C., Rigoux, L., Daunizeau, J. & Pessiglione, M. Neurocomputational account of how the human brain decides when to have a break. Proc. Natl. Acad. Sci. U.S.A. 110, 2641–2646 (2013).

8. Chong, T. T.-J. et al. Neurocomputational mechanisms underlying subjective valuation of effort costs. PLoS Biol 15, e1002598 (2017).

9. Steward, G., Looi, V. & Chib, V. S. The Neurobiology of Cognitive Fatigue and Its Influence on Effort-Based Choice. J. Neurosci. 45, e1612242025 (2025).

10. Westbrook, A., Kester, D. & Braver, T. S. What Is the Subjective Cost of Cognitive Effort? Load, Trait, and Aging Effects Revealed by Economic Preference. PLoS ONE 8, e68210 (2013).

11. Westbrook, A., Lamichhane, B. & Braver, T. The Subjective Value of Cognitive Effort is Encoded by a Domain-General Valuation Network. J. Neurosci. 39, 3934–3947 (2019).

12. Culbreth, A. J. et al. A Transdiagnostic Study of Effort-Cost Decision-Making in Psychotic and Mood Disorders. Schizophrenia Bulletin sbad155 (2023) doi:10.1093/schbul/sbad155.

13. Barch, D. M. et al. Dissociation of Cognitive Effort–Based Decision Making and Its Associations With Symptoms, Cognition, and Everyday Life Function Across Schizophrenia, Bipolar Disorder, and Depression. Biological Psychiatry 94, 501–510 (2023).

14. Moran, E. K., Prevost, C., Culbreth, A. J. & Barch, D. M. Effort-cost decision-making in psychotic and mood disorders. Journal of Psychopathology and Clinical Science 132, 490–498 (2023).

15. Zou, Y. et al. Effort–cost computation in a transdiagnostic psychiatric sample: Differences among patients with schizophrenia, bipolar disorder, and major depressive disorder. Psych J 9, 210–222 (2020).

16. Westbrook, A. et al. Economic Choice and Heart Rate Fractal Scaling Indicate That Cognitive Effort Is Reduced by Depression and Boosted by Sad Mood. Biological Psychiatry: Cognitive Neuroscience and Neuroimaging 8, 687–694 (2023).

17. Hershenberg, R. et al. Diminished effort on a progressive ratio task in both unipolar and bipolar depression. Journal of Affective Disorders 196, 97–100 (2016).

18. Ang, Y.-S., Gelda, S. E. & Pizzagalli, D. A. Cognitive effort-based decision-making in major depressive disorder. Psychol. Med. 1–8 (2022) doi:10.1017/S0033291722000964.

19. Vinckier, F. et al. Elevated Effort Cost Identified by Computational Modeling as a Distinctive Feature Explaining Multiple Behaviors in Patients With Depression. Biological Psychiatry: Cognitive Neuroscience and Neuroimaging 7, 1158–1169 (2022).

20. Horne, S. J., Topp, T. E. & Quigley, L. Depression and the willingness to expend cognitive and physical effort for rewards: A systematic review. Clinical Psychology Review 88, 102065 (2021).

21. Berwian, I. M. et al. Computational Mechanisms of Effort and Reward Decisions in Patients With Depression and Their Association With Relapse After Antidepressant Discontinuation. JAMA Psychiatry 77, 513 (2020).

22. Yang, X. et al. Motivational deficits in effort-based decision making in individuals with subsyndromal depression, first-episode and remitted depression patients. Psychiatry Research 220, 874–882 (2014).

23. Treadway, M. T., Buckholtz, J. W., Schwartzman, A. N., Lambert, W. E. & Zald, D. H. Worth the ‘EEfRT’? The Effort Expenditure for Rewards Task as an Objective Measure of Motivation and Anhedonia. PLoS ONE 4, e6598 (2009).

24. Treadway, M. T., Bossaller, N. A., Shelton, R. C. & Zald, D. H. Effort-based decision-making in major depressive disorder: A translational model of motivational anhedonia. Journal of Abnormal Psychology 121, 553–558 (2012).

25. Yang, X. et al. Diminished caudate and superior temporal gyrus responses to effort-based decision making in patients with first-episode major depressive disorder. Progress in Neuro-Psychopharmacology and Biological Psychiatry 64, 52–59 (2016).

26. Blain, B., Hollard, G. & Pessiglione, M. Neural mechanisms underlying the impact of daylong cognitive work on economic decisions. Proc. Natl. Acad. Sci. U.S.A. 113, 6967–6972 (2016).

27. Blain, B. et al. Neuro-computational Impact of Physical Training Overload on Economic Decision-Making. Current Biology 29, 3289–3297.e4 (2019).

28. Sheehan, D. V. et al. Reliability and Validity of the Mini International Neuropsychiatric Interview for Children and Adolescents (MINI-KID). J. Clin. Psychiatry 71, 313–326 (2010).

29. National Institute of Mental Health Molecular Genetics Initiative. Diagnostic Interview for Genetic Studies (DIGS). (2003).

30. Beck, A. T. & Steer, R. A. Manual for the Beck Depression Inventory. (The Psychological Corporation., San Antonio, TX, 1993).

31. The Consortium of Multiple Sclerosis Centers Health Services Research Subcommittee. Multiple Sclerosis Quality of Life Inventory: A User’s Manual. (National Multiple Sclerosis Societ, New York, NY, 1997).

32. The Mathworks Inc. MATLAB version: 9.14.0 (R2023a). The MathWorks Inc. (2023).

33. Vogel, T. A., Savelson, Z. M., Otto, A. R. & Roy, M. Forced choices reveal a trade-off between cognitive effort and physical pain. eLife 9, e59410 (2020).

34. Massar, S. A. A., Pu, Z., Chen, C. & Chee, M. W. L. Losses Motivate Cognitive Effort More Than Gains in Effort-Based Decision Making and Performance. Front. Hum. Neurosci. 14, 287 (2020).

35. Capanu, M., Gönen, M. & Begg, C. B. An assessment of estimation methods for generalized linear mixed models with binary outcomes. Statist. Med. 32, 4550–4566 (2013).

36. Ju, K., Lin, L., Chu, H., Cheng, L.-L. & Xu, C. Laplace approximation, penalized quasi-likelihood, and adaptive Gauss–Hermite quadrature for generalized linear mixed models: towards meta-analysis of binary outcome with sparse data. BMC Med Res Methodol 20, 152 (2020).

37. R Core Team. R: A Language and Environment for Statistical Computing. (R Foundation for Statistical Computing, Vienna, Austria, 2022).

38. William Revelle. Psych: Procedures for Psychological, Psychometric, and Personality Research. (Northwestern University, Evanston, Illinois, 2023).

39. Raiche, G. & Magis, D. nFactors: Parallel Analysis and Other Non Graphical Solutions to the Cattell Scree Test. (2022).

40. Costello, A. B. & Osborne, J. Best practices in exploratory factor analysis: four recommendations for getting the most from your analysis. doi:10.7275/JYJ1-4868.

41. Watkins, M. W. A Step-by-Step Guide to Exploratory Factor Analysis with R and Rstudio / Marley W. Watkins. A step-by-step guide to exploratory factor analysis with R and Rstudio (Routledge, New York, NY, 2021).

42. Acar GüvendiR, M. & Özer Özkan, Y. Item Removal Strategies Conducted in Exploratory Factor Analysis: A Comparative Study. International Journal of Assessment Tools in Education 9, 165–180 (2022).

43. DiStefano, C., Zhu, M. & Mîndrilã, D. Understanding and Using Factor Scores: Considerations for the Applied Researcher. doi:10.7275/DA8T-4G52.

44. Culbreth, A. J., Moran, E. K. & Barch, D. M. Effort-cost decision-making in psychosis and depression: could a similar behavioral deficit arise from disparate psychological and neural mechanisms? Psychol. Med. 48, 889–904 (2018).

45. Liu, W. et al. Deficits in sustaining reward responses in subsyndromal and syndromal major depression. Progress in Neuro-Psychopharmacology and Biological Psychiatry 35, 1045–1052 (2011).

46. Pizzagalli, D. A. et al. Reduced Caudate and Nucleus Accumbens Response to Rewards in Unmedicated Individuals With Major Depressive Disorder. AJP 166, 702–710 (2009).

47. Sherdell, L., Waugh, C. E. & Gotlib, I. H. Anticipatory pleasure predicts motivation for reward in major depression. Journal of Abnormal Psychology 121, 51–60 (2012).

48. Whitton, A. E., Treadway, M. T. & Pizzagalli, D. A. Reward processing dysfunction in major depression, bipolar disorder and schizophrenia. Current Opinion in Psychiatry 28, 7–12 (2015).

49. Whitton, A. E. et al. Blunted Neural Responses to Reward in Remitted Major Depression: A High-Density Event-Related Potential Study. Biological Psychiatry: Cognitive Neuroscience and Neuroimaging 1, 87–95 (2016).

50. Grahek, I., Shenhav, A., Musslick, S., Krebs, R. M. & Koster, E. H. W. Motivation and cognitive control in depression. Neuroscience & Biobehavioral Reviews 102, 371–381 (2019).

51. Culbreth, A. J., Dershwitz, S. D., Barch, D. M. & Moran, E. K. Associations Between Cognitive and Physical Effort–Based Decision Making in People With Schizophrenia and Healthy Control Subjects. Biological Psychiatry: Cognitive Neuroscience and Neuroimaging 8, 695–702 (2023).

