## Supplementary Information for "Fatigue Symptoms Influence Effort-Based Decision-Making in Major Depressive Disorder"

Vikram S. Chib

716 North Broadway

Rm 241

Baltimore, MD 21205, USA

443-923-2716

#### Methods

##### Factor Analysis 1: All Items

Factor Analysis 1 (FA1) was a data retention conservative method with an item cross-correlation threshold of 0.9, resulting in no items removed from the original factor analysis due to issues of multicollinearity. All items were retained for factor analysis to retain the maximum amount of data. Tests on the full data set included Bartlett's test of sphericity($\chi2(861) = 2647.6$, $P = 1.09\text{-}180$) and Kaiser-Meyer-Olkin factor adequacy (Overall Measure of Sampling Adequacy (MSA) = 0.63, lowest item MSA = 0.38). The full original dataset was not satisfactory for exploratory factor analysis (EFA), but was used to show the stability of the structure and predictive power of the factor scores, even when the conditions of EFA were less than ideal. The factor analysis from this point on was conducted using the same software and functions detailed in the main text methods.

##### Factor Analysis 2: Removing Items with Cross-Correlation > 0.8 and Cross-Factor Loadings

Factor Analysis 2 (FA2), as detailed in label tables, is the factor analysis detailed in the methods of the main text.

##### Factor Analysis 3: Removing Items with Cross-Correlation > 0.8 and KMO MSA < 0.6

Factor Analys 3 (FA3) was based on a method to remove items with low individual KMO MSAs prior to a preliminary factor analysis. The same items in FA 2 were removed due to high cross-correlation and potential contribution to multicollinearity (BDI Items 4 and 15, and MFIS Items 15,19, 21). Additionally, items with a KMO MSA < 0.6 were removed (BDI Items 18-21). The resulted in a satisfactory overall KMO MSA of 0.81, with the lowest score of an individual item being 0.63. Beyond the point of item removal, the factor analysis was conducted in the same manner as the one described in the main text methods. -

#### Figures

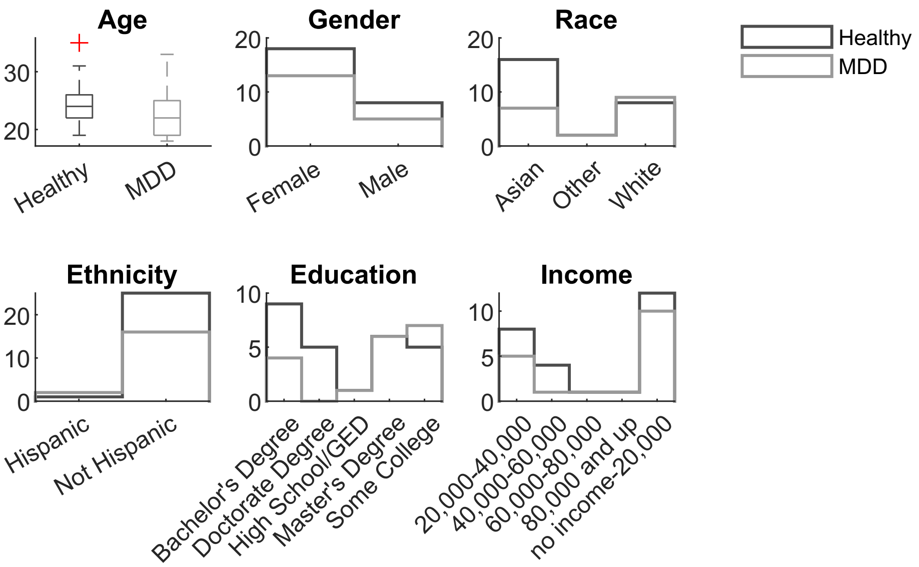

***Figure S1.*** *Demographic characteristics of the healthy and MDD cohorts.*

### Tables

***Table S1. Counts of participants on psychotropic medications in the MDD Cohort.*** *No persons in the healthy cohort reported a history of psychotropic medications.*

| ***Medication*** | ***Participant Count*** |
|---|---|
| bupropion | 5 |
| buspirone | 1 |
| escitalopram | 6 |
| gabapentin | 1 |
| none | 4 |
| paroxetine | 1 |
| sertraline | 2 |
| venlafaxine | 3 |

***Table S2. Between Cohort Model.***

| *Predictors* | *Estimate* | *SE* | *tStat* | *DF* | *P Value* |
|---|---|---|---|---|---|
| *Effort*$^2$ | -4.175 | 0.288 | -14.480 | 1039 | 2.029 E -43 |
| *Reward* | 4.700 | 0.341 | 13.786 | 1039 | 7.896 E -40 |
| *Cohort MDD* | -0.751 | 0.468 | -1.604 | 1039 | 0.109 |

***Table S3. Between Cohort Model with Education Profiled Out.***

| *Predictors* | *Estimate* | *SE* | *tStat* | *DF* | *P Value* |
|---|---|---|---|---|---|
| *Effort*$^2$ | -4.186 | 0.290 | -14.435 | 1038 | 3.510 E -43 |
| *Reward* | 4.675 | 0.343 | 13.635 | 1038 | 4.604 E -39 |
| *Cohort MDD* | -0.876 | 0.537 | -1.631 | 1038 | 0.103 |
| *Years of Education* | 0.286 | 0.603 | 0.474 | 1038 | 0.635 |

***Table S4. Paradigm Choice Set.*** *Unique choices presented to participants. Participants would choose whether to perform one of the offers below for the stated effort and reward, or to perform the default 1-back task for a $1 reward.*

| Effort Level (*n*) | Reward ($) |
|---|---|
| 2 | 1 |
| 3 | 1 |
| 4 | 1 |
| 2 | 2 |
| 3 | 2 |
| 4 | 2 |
| 2 | 3 |
| 3 | 3 |
| 4 | 3 |
| 2 | 4 |
| 3 | 4 |
| 4 | 4 |
| 2 | 5 |
| 3 | 5 |
| 4 | 5 |
| 2 | 6 |
| 3 | 6 |
| 4 | 6 |
| 2 | 7 |
| 3 | 7 |
| 4 | 7 |
| 2 | 8 |
| 3 | 8 |
| 4 | 8 |

***Table S5. No Significant Difference in n-back Performance Between Cohorts.*** *N-Way ANOVA Type III Sum of Squares*

| | Sum of Squares | DF | Mean Squares | F | P Value |
|---|---|---|---|---|---|
| *n-back Level* | 0.683 | 3 | 0.228 | 23.48 | 3.414 E -12 |
| *Cohort* | 0.001 | 1 | 0.001 | 0.12 | 0.730 |
| *Participant (Nested in Cohort)* | 0.779 | 42 | 0.019 | 1.91 | 0.003 |
| *Error* | 1.251 | 129 | 0.010 | | |
| *Total* | 2.715 | 175 | | | |

***Table S6. No Significant Difference in n-back Difficulty Perception Between Cohorts.*** *N-Way ANOVA Type III Sum of Squares*

| | Sum of Squares | DF | Mean Squares | F | P Value |
|---|---|---|---|---|---|
| *n-back Level* | 220.16 | 3 | 73.388 | 33.23 | 5.569 E -16 |
| *Cohort* | 1.97 | 1 | 1.968 | 0.89 | 0.347 |
| *Participant (Nested in Cohort)* | 65.35 | 42 | 1.556 | 0.70 | 0.90 |
| *Error* | 284.91 | 129 | 2.208 | | |
| *Total* | 572.39 | 175 | | | |

***Table S7. n-back Difficulty Perception Not Significantly Affected by Depression or Fatigue Factors (Healthy and MDD Cohort).*** *Generalized Linear Mixed Effects Model with Random Intercept by participant ID*

| ***Model*** | ***Vars*** | ***Estimate*** | ***P Value*** |
|---|---|---|---|
| *Depression Factor Model* | Intercept | 1.62 | 4.85 E-09 |
| | *n*-back Level | 0.98 | 1.78 E-19 |
| | Depression Factor | 0.08 | 0.480 |
| *Fatigue Factor Model* | Intercept | 1.62 | 4.55 E-09 |
| | *n*-back Level | 0.98 | 1.50 E-19 |
| | Fatigue Factor | 0.13 | 0.235 |

***Table S8. n-back Difficulty Perception Is Better Predicted by n-back Level than Task Accuracy.*** *Generalized Linear Mixed Effects Model with Random Intercept by participant ID*

| Model | Vars | Estimate | P Value | AIC |
|---|---|---|---|---|
| *Task Accuracy Model* | Intercept | 3.99 | 4.97E-58 | 690 |
| | Task Accuracy | -0.83 | 1.04E-06 | |
| | MDD Cohort | 0.22 | 0.399 | |
| | Task Accuracy:MDD Cohort | 0.38 | 0.138 | |
| *Effort Level Model* | Intercept | 3.99 | 2.18E-67 | 635 |
| | *n*-back Level | 1.01 | 1.57E-11 | |
| | MDD Cohort | 0.22 | 0.325 | |
| | *n*-back Level:MDD Cohort | 0.23 | 0.294 | |

***Table S9. Mean Task Accuracy Does Not Meaningfully Add to Models of Choice.*** *Generalized Linear Mixed Effects Model with Random Intercept by participant ID*

| Variables | Estimate | P Value |
|---|---|---|
| *(n-back Level)$^2$* | -4.14 | 1.76E-42 |
| *Reward* | 4.77 | 3.71E-38 |
| *Mean Accuracy* | -0.36 | 0.417 |
| *MDD Cohort* | -0.48 | 0.402 |

***Table S10. n-back Level Is the Best Model of Effort Cost in Choice.*** *Generalized Linear Mixed Effects Model with Random Intercept by participant ID. Choice ~ Effort Cost + Reward + Cohort + (1 | participant)*

| Effort Cost Variables | AIC |
|---|---|
| *(n-back Level)$^2$* | 856 |
| *Difficulty Rating* | 1079 |
| *Task Accuracy* | 1169 |

***Table S11. Scale item correlations.*** *Showing the correlation of all items and those dropped for overcorrelation.*

| | | 1 | 2 | 3 | 4 | 5 | 6 | 7 | 8 | 9 | 10 | 11 | 12 | 13 | 14 | 15 | 16 | 17 | 18 | 19 | 20 | 21 | 1 | 2 | 3 | 4 | 5 | 6 | 7 | 8 | 9 | 10 | 11 | 12 | 13 | 14 | 15 | 16 | 17 | 18 | 19 | 20 | 21 |
|---|---|---|---|---|---|---|---|---|---|---|---|---|---|---|---|---|---|---|---|---|---|---|---|---|---|---|---|---|---|---|---|---|---|---|---|---|---|---|---|---|---|---|---|
| BDI | 1 | 1.00 | -- | -- | -- | -- | -- | -- | -- | -- | -- | -- | -- | -- | -- | -- | -- | -- | -- | -- | -- | -- | -- | -- | -- | -- | -- | -- | -- | -- | -- | -- | -- | -- | -- | -- | -- | -- | -- | -- | -- | -- | -- |
| | 2 | 0.74 | 1.00 | -- | -- | -- | -- | -- | -- | -- | -- | -- | -- | -- | -- | -- | -- | -- | -- | -- | -- | -- | -- | -- | -- | -- | -- | -- | -- | -- | -- | -- | -- | -- | -- | -- | -- | -- | -- | -- | -- | -- | -- |
| | 3 | 0.60 | 0.58 | 1.00 | -- | -- | -- | -- | -- | -- | -- | -- | -- | -- | -- | -- | -- | -- | -- | -- | -- | -- | -- | -- | -- | -- | -- | -- | -- | -- | -- | -- | -- | -- | -- | -- | -- | -- | -- | -- | -- | -- | -- |
| | 4 | 0.85 | 0.68 | 0.55 | 1.00 | -- | -- | -- | -- | -- | -- | -- | -- | -- | -- | -- | -- | -- | -- | -- | -- | -- | -- | -- | -- | -- | -- | -- | -- | -- | -- | -- | -- | -- | -- | -- | -- | -- | -- | -- | -- | -- | -- |
| | 5 | 0.70 | 0.68 | 0.65 | 0.64 | 1.00 | -- | -- | -- | -- | -- | -- | -- | -- | -- | -- | -- | -- | -- | -- | -- | -- | -- | -- | -- | -- | -- | -- | -- | -- | -- | -- | -- | -- | -- | -- | -- | -- | -- | -- | -- | -- | -- |
| | 6 | 0.64 | 0.70 | 0.55 | 0.48 | 0.59 | 1.00 | -- | -- | -- | -- | -- | -- | -- | -- | -- | -- | -- | -- | -- | -- | -- | -- | -- | -- | -- | -- | -- | -- | -- | -- | -- | -- | -- | -- | -- | -- | -- | -- | -- | -- | -- | -- |
| | 7 | 0.77 | 0.75 | 0.66 | 0.77 | 0.71 | 0.67 | 1.00 | -- | -- | -- | -- | -- | -- | -- | -- | -- | -- | -- | -- | -- | -- | -- | -- | -- | -- | -- | -- | -- | -- | -- | -- | -- | -- | -- | -- | -- | -- | -- | -- | -- | -- | -- |
| | 8 | 0.59 | 0.61 | 0.75 | 0.58 | 0.75 | 0.59 | 0.77 | 1.00 | -- | -- | -- | -- | -- | -- | -- | -- | -- | -- | -- | -- | -- | -- | -- | -- | -- | -- | -- | -- | -- | -- | -- | -- | -- | -- | -- | -- | -- | -- | -- | -- | -- | -- |
| | 9 | 0.74 | 0.78 | 0.45 | 0.56 | 0.58 | 0.76 | 0.61 | 0.59 | 1.00 | -- | -- | -- | -- | -- | -- | -- | -- | -- | -- | -- | -- | -- | -- | -- | -- | -- | -- | -- | -- | -- | -- | -- | -- | -- | -- | -- | -- | -- | -- | -- | -- | -- |
| | 10 | 0.74 | 0.60 | 0.44 | 0.72 | 0.67 | 0.53 | 0.55 | 0.51 | 0.69 | 1.00 | -- | -- | -- | -- | -- | -- | -- | -- | -- | -- | -- | -- | -- | -- | -- | -- | -- | -- | -- | -- | -- | -- | -- | -- | -- | -- | -- | -- | -- | -- | -- | -- |
| | 11 | 0.76 | 0.73 | 0.58 | 0.68 | 0.64 | 0.51 | 0.77 | 0.64 | 0.65 | 0.61 | 1.00 | -- | -- | -- | -- | -- | -- | -- | -- | -- | -- | -- | -- | -- | -- | -- | -- | -- | -- | -- | -- | -- | -- | -- | -- | -- | -- | -- | -- | -- | -- | -- |
| | 12 | 0.80 | 0.62 | 0.51 | 0.76 | 0.64 | 0.59 | 0.64 | 0.58 | 0.62 | 0.74 | 0.68 | 1.00 | -- | -- | -- | -- | -- | -- | -- | -- | -- | -- | -- | -- | -- | -- | -- | -- | -- | -- | -- | -- | -- | -- | -- | -- | -- | -- | -- | -- | -- | -- |
| | 13 | 0.59 | 0.58 | 0.56 | 0.70 | 0.76 | 0.40 | 0.69 | 0.77 | 0.46 | 0.61 | 0.53 | 0.57 | 1.00 | -- | -- | -- | -- | -- | -- | -- | -- | -- | -- | -- | -- | -- | -- | -- | -- | -- | -- | -- | -- | -- | -- | -- | -- | -- | -- | -- | -- | -- |
| | 14 | 0.60 | 0.41 | 0.62 | 0.56 | 0.68 | 0.43 | 0.67 | 0.67 | 0.35 | 0.45 | 0.51 | 0.58 | 0.58 | 1.00 | -- | -- | -- | -- | -- | -- | -- | -- | -- | -- | -- | -- | -- | -- | -- | -- | -- | -- | -- | -- | -- | -- | -- | -- | -- | -- | -- | -- |
| | 15 | 0.80 | 0.67 | 0.56 | 0.83 | 0.67 | 0.51 | 0.81 | 0.72 | 0.62 | 0.75 | 0.67 | 0.70 | 0.76 | 0.56 | 1.00 | -- | -- | -- | -- | -- | -- | -- | -- | -- | -- | -- | -- | -- | -- | -- | -- | -- | -- | -- | -- | -- | -- | -- | -- | -- | -- | -- |
| | 16 | 0.60 | 0.64 | 0.38 | 0.58 | 0.44 | 0.37 | 0.63 | 0.56 | 0.57 | 0.53 | 0.58 | 0.58 | 0.59 | 0.34 | 0.62 | 1.00 | -- | -- | -- | -- | -- | -- | -- | -- | -- | -- | -- | -- | -- | -- | -- | -- | -- | -- | -- | -- | -- | -- | -- | -- | -- | -- |
| | 17 | 0.73 | 0.67 | 0.49 | 0.71 | 0.68 | 0.51 | 0.78 | 0.67 | 0.62 | 0.58 | 0.67 | 0.62 | 0.68 | 0.58 | 0.73 | 0.70 | 1.00 | -- | -- | -- | -- | -- | -- | -- | -- | -- | -- | -- | -- | -- | -- | -- | -- | -- | -- | -- | -- | -- | -- | -- | -- | -- |
| | 18 | 0.62 | 0.61 | 0.30 | 0.60 | 0.52 | 0.36 | 0.50 | 0.40 | 0.53 | 0.51 | 0.39 | 0.52 | 0.45 | 0.43 | 0.55 | 0.52 | 0.52 | 1.00 | -- | -- | -- | -- | -- | -- | -- | -- | -- | -- | -- | -- | -- | -- | -- | -- | -- | -- | -- | -- | -- | -- | -- | -- |
| | 19 | 0.38 | 0.60 | 0.25 | 0.41 | 0.39 | 0.38 | 0.45 | 0.37 | 0.47 | 0.41 | 0.36 | 0.45 | 0.38 | 0.17 | 0.42 | 0.59 | 0.50 | 0.62 | 1.00 | -- | -- | -- | -- | -- | -- | -- | -- | -- | -- | -- | -- | -- | -- | -- | -- | -- | -- | -- | -- | -- | -- | -- |
| | 20 | 0.42 | 0.50 | 0.33 | 0.35 | 0.38 | 0.38 | 0.52 | 0.48 | 0.41 | 0.34 | 0.29 | 0.40 | 0.33 | 0.54 | 0.47 | 0.53 | 0.58 | 0.26 | 0.45 | 1.00 | -- | -- | -- | -- | -- | -- | -- | -- | -- | -- | -- | -- | -- | -- | -- | -- | -- | -- | -- | -- | -- | -- |
| | 21 | 0.42 | 0.33 | -0.01 | 0.37 | 0.34 | 0.32 | 0.32 | 0.13 | 0.22 | 0.25 | 0.31 | 0.46 | 0.19 | 0.34 | 0.28 | 0.35 | 0.46 | 0.35 | 0.33 | 0.35 | 1.00 | -- | -- | -- | -- | -- | -- | -- | -- | -- | -- | -- | -- | -- | -- | -- | -- | -- | -- | -- | -- | -- |
| MFIS | 1 | 0.47 | 0.47 | 0.35 | 0.54 | 0.52 | 0.46 | 0.68 | 0.57 | 0.44 | 0.50 | 0.51 | 0.35 | 0.49 | 0.43 | 0.62 | 0.40 | 0.60 | 0.28 | 0.31 | 0.39 | 0.19 | 1.00 | -- | -- | -- | -- | -- | -- | -- | -- | -- | -- | -- | -- | -- | -- | -- | -- | -- | -- | -- | -- |
| | 2 | 0.51 | 0.53 | 0.46 | 0.58 | 0.68 | 0.55 | 0.66 | 0.61 | 0.47 | 0.55 | 0.50 | 0.49 | 0.59 | 0.54 | 0.64 | 0.36 | 0.67 | 0.32 | 0.27 | 0.37 | 0.32 | 0.78 | 1.00 | -- | -- | -- | -- | -- | -- | -- | -- | -- | -- | -- | -- | -- | -- | -- | -- | -- | -- | -- |
| | 3 | 0.61 | 0.59 | 0.43 | 0.63 | 0.63 | 0.68 | 0.73 | 0.62 | 0.57 | 0.52 | 0.62 | 0.58 | 0.49 | 0.46 | 0.65 | 0.40 | 0.60 | 0.32 | 0.31 | 0.33 | 0.36 | 0.80 | 0.78 | 1.00 | -- | -- | -- | -- | -- | -- | -- | -- | -- | -- | -- | -- | -- | -- | -- | -- | -- | -- |
| | 4 | 0.50 | 0.43 | 0.34 | 0.59 | 0.59 | 0.41 | 0.67 | 0.58 | 0.44 | 0.47 | 0.42 | 0.40 | 0.65 | 0.40 | 0.65 | 0.42 | 0.71 | 0.27 | 0.20 | 0.36 | 0.20 | 0.69 | 0.74 | 0.66 | 1.00 | -- | -- | -- | -- | -- | -- | -- | -- | -- | -- | -- | -- | -- | -- | -- | -- | -- |
| | 5 | 0.63 | 0.68 | 0.48 | 0.64 | 0.66 | 0.60 | 0.73 | 0.75 | 0.67 | 0.52 | 0.64 | 0.61 | 0.70 | 0.58 | 0.65 | 0.55 | 0.66 | 0.38 | 0.36 | 0.50 | 0.29 | 0.64 | 0.65 | 0.71 | 0.60 | 1.00 | -- | -- | -- | -- | -- | -- | -- | -- | -- | -- | -- | -- | -- | -- | -- | -- |
| | 6 | 0.43 | 0.40 | 0.37 | 0.50 | 0.58 | 0.51 | 0.50 | 0.45 | 0.32 | 0.51 | 0.34 | 0.48 | 0.59 | 0.47 | 0.45 | 0.42 | 0.46 | 0.34 | 0.32 | 0.24 | 0.21 | 0.40 | 0.59 | 0.46 | 0.46 | 0.42 | 1.00 | -- | -- | -- | -- | -- | -- | -- | -- | -- | -- | -- | -- | -- | -- | -- |
| | 7 | 0.63 | 0.61 | 0.53 | 0.66 | 0.74 | 0.55 | 0.70 | 0.68 | 0.58 | 0.62 | 0.58 | 0.51 | 0.69 | 0.63 | 0.65 | 0.52 | 0.74 | 0.47 | 0.34 | 0.45 | 0.17 | 0.64 | 0.72 | 0.64 | 0.66 | 0.64 | 0.69 | 1.00 | -- | -- | -- | -- | -- | -- | -- | -- | -- | -- | -- | -- | -- | -- |
| | 8 | 0.64 | 0.48 | 0.47 | 0.68 | 0.53 | 0.46 | 0.66 | 0.49 | 0.46 | 0.58 | 0.56 | 0.62 | 0.57 | 0.51 | 0.59 | 0.58 | 0.64 | 0.35 | 0.36 | 0.37 | 0.32 | 0.65 | 0.67 | 0.62 | 0.66 | 0.61 | 0.52 | 0.67 | 1.00 | -- | -- | -- | -- | -- | -- | -- | -- | -- | -- | -- | -- | -- |
| | 9 | 0.72 | 0.63 | 0.41 | 0.69 | 0.63 | 0.59 | 0.71 | 0.58 | 0.57 | 0.66 | 0.63 | 0.65 | 0.66 | 0.42 | 0.74 | 0.51 | 0.71 | 0.31 | 0.23 | 0.39 | 0.33 | 0.61 | 0.69 | 0.69 | 0.73 | 0.65 | 0.52 | 0.66 | 0.72 | 1.00 | -- | -- | -- | -- | -- | -- | -- | -- | -- | -- | -- | -- |
| | 10 | 0.49 | 0.49 | 0.36 | 0.45 | 0.65 | 0.67 | 0.62 | 0.55 | 0.63 | 0.51 | 0.43 | 0.41 | 0.55 | 0.51 | 0.52 | 0.38 | 0.63 | 0.29 | 0.25 | 0.39 | 0.23 | 0.60 | 0.68 | 0.63 | 0.61 | 0.63 | 0.61 | 0.79 | 0.56 | 0.57 | 1.00 | -- | -- | -- | -- | -- | -- | -- | -- | -- | -- | -- |
| | 11 | 0.58 | 0.64 | 0.52 | 0.68 | 0.76 | 0.44 | 0.72 | 0.75 | 0.50 | 0.57 | 0.59 | 0.55 | 0.78 | 0.51 | 0.70 | 0.58 | 0.70 | 0.51 | 0.35 | 0.32 | 0.26 | 0.56 | 0.65 | 0.64 | 0.64 | 0.69 | 0.57 | 0.72 | 0.55 | 0.66 | 0.63 | 1.00 | -- | -- | -- | -- | -- | -- | -- | -- | -- | -- |
| | 12 | 0.68 | 0.65 | 0.40 | 0.72 | 0.61 | 0.54 | 0.71 | 0.57 | 0.59 | 0.59 | 0.59 | 0.58 | 0.67 | 0.50 | 0.68 | 0.52 | 0.70 | 0.40 | 0.32 | 0.39 | 0.38 | 0.70 | 0.80 | 0.75 | 0.71 | 0.68 | 0.48 | 0.71 | 0.71 | 0.74 | 0.64 | 0.67 | 1.00 | -- | -- | -- | -- | -- | -- | -- | -- | -- |
| | 13 | 0.56 | 0.56 | 0.17 | 0.53 | 0.59 | 0.50 | 0.55 | 0.47 | 0.53 | 0.47 | 0.50 | 0.48 | 0.51 | 0.42 | 0.49 | 0.49 | 0.65 | 0.43 | 0.53 | 0.46 | 0.50 | 0.58 | 0.57 | 0.59 | 0.47 | 0.57 | 0.49 | 0.68 | 0.59 | 0.53 | 0.68 | 0.54 | 0.66 | 1.00 | -- | -- | -- | -- | -- | -- | -- | -- |
| | 14 | 0.65 | 0.64 | 0.37 | 0.61 | 0.67 | 0.62 | 0.71 | 0.54 | 0.67 | 0.52 | 0.62 | 0.57 | 0.53 | 0.53 | 0.63 | 0.51 | 0.70 | 0.34 | 0.35 | 0.49 | 0.38 | 0.65 | 0.70 | 0.73 | 0.65 | 0.63 | 0.54 | 0.69 | 0.62 | 0.68 | 0.77 | 0.59 | 0.69 | 0.73 | 1.00 | -- | -- | -- | -- | -- | -- | -- |
| | 15 | 0.65 | 0.59 | 0.38 | 0.74 | 0.57 | 0.52 | 0.68 | 0.59 | 0.51 | 0.59 | 0.61 | 0.68 | 0.65 | 0.51 | 0.67 | 0.54 | 0.65 | 0.34 | 0.23 | 0.39 | 0.46 | 0.64 | 0.76 | 0.74 | 0.66 | 0.66 | 0.50 | 0.65 | 0.70 | 0.80 | 0.54 | 0.70 | 0.89 | 0.60 | 0.67 | 1.00 | -- | -- | -- | -- | -- | -- |
| | 16 | 0.65 | 0.64 | 0.42 | 0.72 | 0.62 | 0.41 | 0.75 | 0.65 | 0.50 | 0.47 | 0.66 | 0.58 | 0.69 | 0.53 | 0.70 | 0.62 | 0.70 | 0.35 | 0.20 | 0.41 | 0.42 | 0.64 | 0.65 | 0.72 | 0.67 | 0.75 | 0.40 | 0.64 | 0.62 | 0.72 | 0.55 | 0.77 | 0.80 | 0.56 | 0.70 | 0.81 | 1.00 | -- | -- | -- | -- | -- |
| | 17 | 0.63 | 0.65 | 0.40 | 0.71 | 0.52 | 0.51 | 0.69 | 0.54 | 0.59 | 0.54 | 0.62 | 0.55 | 0.66 | 0.49 | 0.61 | 0.66 | 0.59 | 0.43 | 0.35 | 0.38 | 0.25 | 0.47 | 0.51 | 0.58 | 0.52 | 0.62 | 0.54 | 0.77 | 0.70 | 0.64 | 0.59 | 0.58 | 0.71 | 0.63 | 0.67 | 0.73 | 0.68 | 1.00 | -- | -- | -- | -- |
| | 18 | 0.66 | 0.60 | 0.39 | 0.75 | 0.50 | 0.54 | 0.66 | 0.59 | 0.59 | 0.51 | 0.53 | 0.73 | 0.62 | 0.53 | 0.66 | 0.66 | 0.69 | 0.45 | 0.41 | 0.45 | 0.50 | 0.57 | 0.65 | 0.71 | 0.58 | 0.74 | 0.46 | 0.56 | 0.69 | 0.66 | 0.51 | 0.64 | 0.78 | 0.57 | 0.69 | 0.85 | 0.78 | 0.69 | 1.00 | -- | -- | -- |
| | 19 | 0.65 | 0.62 | 0.40 | 0.69 | 0.72 | 0.54 | 0.65 | 0.67 | 0.58 | 0.64 | 0.59 | 0.64 | 0.70 | 0.53 | 0.68 | 0.58 | 0.80 | 0.46 | 0.39 | 0.45 | 0.45 | 0.72 | 0.84 | 0.75 | 0.74 | 0.73 | 0.56 | 0.75 | 0.70 | 0.75 | 0.65 | 0.74 | 0.82 | 0.75 | 0.72 | 0.83 | 0.73 | 0.62 | 0.78 | 1.00 | -- | -- |
| | 20 | 0.59 | 0.54 | 0.31 | 0.62 | 0.52 | 0.36 | 0.52 | 0.43 | 0.52 | 0.55 | 0.57 | 0.54 | 0.46 | 0.54 | 0.48 | 0.55 | 0.57 | 0.50 | 0.45 | 0.47 | 0.28 | 0.49 | 0.47 | 0.50 | 0.35 | 0.54 | 0.46 | 0.74 | 0.63 | 0.45 | 0.54 | 0.47 | 0.59 | 0.70 | 0.60 | 0.57 | 0.51 | 0.72 | 0.59 | 0.61 | 1.00 | -- |
| | 21 | 0.73 | 0.69 | 0.43 | 0.68 | 0.61 | 0.60 | 0.72 | 0.61 | 0.70 | 0.56 | 0.64 | 0.62 | 0.52 | 0.50 | 0.65 | 0.61 | 0.83 | 0.53 | 0.45 | 0.47 | 0.42 | 0.62 | 0.64 | 0.70 | 0.65 | 0.64 | 0.36 | 0.70 | 0.66 | 0.65 | 0.65 | 0.64 | 0.71 | 0.69 | 0.72 | 0.69 | 0.70 | 0.64 | 0.74 | 0.78 | 0.62 | 1.00 |

*Table S12. Factor loadings for Factor Analysis*

| Item | Description | FA 1. All Items | | FA 2. Controlling for Multicolineariy and Cross-factor loadings | | FA 3. Controlling for Multicolineariy and Poor KMO Scores | |
|---|---|---|---|---|---|---|---|
| | | Fatigue | Depress Mood | Fatigue | Depress Mood | Fatigue | Depress Mood |
| **BDI 1** | Sadness | -0.10 | 1.00 | -0.01 | 0.90 | -0.15 | 1.03 |
| **BDI 2** | Discouraged about Future | -0.02 | 0.86 | -0.01 | 0.88 | -0.06 | 0.89 |
| **BDI 3** | Feelings of Failure | -0.08 | 0.71 | 0.09 | 0.53 | -0.09 | 0.73 |
| **BDI 4** | Anhedonia/Activity Satisfaction | 0.15 | 0.75 | | | | |
| **BDI 5** | Feelings of Guilt | 0.34 | 0.51 | | | 0.34 | 0.52 |
| **BDI 6** | Perception of Generalized Punishment | 0.20 | 0.53 | 0.21 | 0.55 | 0.13 | 0.60 |
| **BDI 7** | Self-Disappointment | 0.31 | 0.61 | | | 0.35 | 0.57 |
| **BDI 8** | Self-Blame | 0.34 | 0.47 | | | 0.32 | 0.50 |
| **BDI 9** | Thoughts of Suicide | -0.02 | 0.79 | -0.02 | 0.84 | -0.08 | 0.86 |
| **BDI 10** | Crying Frequency | 0.07 | 0.71 | 0.12 | 0.68 | 0.04 | 0.72 |
| **BDI 11** | Irritation | 0.01 | 0.80 | 0.08 | 0.74 | -0.08 | 0.90 |
| **BDI 12** | Social Interest | -0.07 | 0.88 | -0.06 | 0.87 | -0.18 | 0.97 |
| **BDI 13** | Decision Making Ability | 0.42 | 0.40 | 0.48 | 0.33 | 0.43 | 0.38 |
| **BDI 14** | Self-Esteem in Appearance | 0.24 | 0.45 | | | 0.25 | 0.45 |
| **BDI 15** | Self-Motivation to Work | 0.22 | 0.68 | | | | |
| **BDI 16** | Insomnia | 0.00 | 0.73 | -0.03 | 0.76 | -0.03 | 0.74 |
| **BDI 17** | Fatiguability | 0.43 | 0.48 | | | 0.43 | 0.46 |
| **BDI 18** | Decreased Appetite | -0.29 | 0.89 | -0.26 | 0.86 | | |
| **BDI 19** | Weight Loss | -0.20 | 0.70 | -0.23 | 0.75 | | |
| **BDI 20** | Physical Health Anxiety | 0.17 | 0.38 | 0.15 | 0.40 | | |
| **BDI 21** | Interest in Sex | 0.17 | 0.28 | 0.05 | 0.39 | | |
| **MFIS 1** | Less Alert | 0.97 | -0.21 | 0.98 | -0.21 | 1.03 | -0.26 |
| **MFIS 2** | Difficulty Paying Attention | 1.07 | -0.23 | 1.03 | -0.19 | 1.06 | -0.22 |
| **MFIS 3** | Difficulty Thinking | 0.81 | 0.04 | 0.80 | 0.06 | 0.78 | 0.08 |
| **MFIS 4** | Clumsiness | 0.95 | -0.17 | 0.99 | -0.22 | 1.00 | -0.22 |
| **MFIS 5** | Forgetfulness | 0.51 | 0.34 | 0.51 | 0.36 | 0.47 | 0.40 |
| **MFIS 6** | Physical Exertion Capacity | 0.51 | 0.12 | 0.52 | 0.13 | 0.54 | 0.10 |
| **MFIS 7** | Physical Effort Motivation | 0.63 | 0.24 | 0.66 | 0.22 | 0.66 | 0.22 |
| **MFIS 8** | Social Motivation | 0.61 | 0.21 | 0.62 | 0.21 | 0.60 | 0.22 |

| | | | | | | | |
|---|---|---|---|---|---|---|---|
| **MFIS 9** | Ability to Leave Home | 0.61 | 0.27 | 0.61 | 0.26 | 0.53 | 0.34 |
| **MFIS 10** | Physical endurance | 0.76 | 0.00 | 0.76 | 0.02 | 0.79 | -0.01 |
| **MFIS 11** | Decision Making Ability | 0.55 | 0.30 | 0.57 | 0.26 | 0.56 | 0.29 |
| **MFIS 12** | Cognitive Effort Motivation | 0.83 | 0.08 | 0.77 | 0.15 | 0.77 | 0.13 |
| **MFIS 13** | Muscle Weakness | 0.58 | 0.18 | 0.47 | 0.30 | 0.55 | 0.20 |
| **MFIS 14** | Physical Discomfort | 0.64 | 0.22 | 0.63 | 0.25 | 0.62 | 0.25 |
| **MFIS 15** | Difficulty Finishing Cognitive Tasks | 0.80 | 0.10 | | | | |
| **MFIS 16** | Executive Function and Organization | 0.66 | 0.22 | 0.66 | 0.21 | 0.60 | 0.27 |
| **MFIS 17** | Physical Task Completion | 0.36 | 0.46 | 0.35 | 0.49 | 0.32 | 0.50 |
| **MFIS 18** | Slow Thinking | 0.56 | 0.31 | | | 0.47 | 0.38 |
| **MFIS 19** | Concentration | 0.86 | 0.07 | | | | |
| **MFIS 20** | Physical Activity Limitation | 0.26 | 0.46 | 0.22 | 0.52 | 0.25 | 0.47 |
| **MFIS 21** | Increased Need of Rest | 0.45 | 0.44 | | | | |

*Table S13. KMO Scores for scale items*

| **Item** | **FA 1** | **FA2** | **FA 3** |
|---|---|---|---|
| BDI 1 | 0.60 | 0.78 | 0.63 |
| BDI 2 | 0.59 | 0.78 | 0.63 |
| BDI 3 | 0.42 | 0.57 | 0.63 |
| BDI 4 | 0.69 | | |
| BDI 5 | 0.80 | | 0.89 |
| BDI 6 | 0.75 | 0.73 | 0.81 |
| BDI 7 | 0.85 | | 0.85 |
| BDI 8 | 0.90 | | 0.70 |
| BDI 9 | 0.50 | 0.65 | 0.72 |
| BDI 10 | 0.48 | 0.63 | 0.78 |
| BDI 11 | 0.75 | 0.84 | 0.87 |
| BDI 12 | 0.59 | 0.79 | 0.79 |
| BDI 13 | 0.59 | 0.68 | 0.83 |
| BDI 14 | 0.62 | | 0.77 |
| BDI 15 | 0.60 | | |
| BDI 16 | 0.64 | 0.71 | 0.80 |
| BDI 17 | 0.89 | | 0.82 |
| BDI 18 | 0.38 | 0.58 | |
| BDI 19 | 0.53 | 0.71 | |
| BDI 20 | 0.40 | 0.59 | |
| BDI 21 | 0.45 | 0.68 | |
| MFIS 1 | 0.49 | 0.71 | 0.77 |
| MFIS 2 | 0.50 | 0.79 | 0.86 |
| MFIS 3 | 0.83 | 0.87 | 0.94 |
| MFIS 4 | 0.68 | 0.88 | 0.87 |
| MFIS 5 | 0.49 | 0.66 | 0.91 |
| MFIS 6 | 0.91 | 0.73 | 0.80 |
| MFIS 7 | 0.62 | 0.81 | 0.79 |
| MFIS 8 | 0.79 | 0.74 | 0.76 |
| MFIS 9 | 0.88 | 0.78 | 0.84 |
| MFIS 10 | 0.64 | 0.86 | 0.80 |
| MFIS 11 | 0.56 | 0.83 | 0.80 |
| MFIS 12 | 0.59 | 0.73 | 0.88 |
| MFIS 13 | 0.67 | 0.90 | 0.72 |
| MFIS 14 | 0.50 | 0.69 | 0.84 |
| MFIS 15 | 0.56 | | |
| MFIS 16 | 0.85 | 0.90 | 0.91 |
| MFIS 17 | 0.62 | 0.79 | 0.77 |
| MFIS 18 | 0.59 | | 0.78 |
| MFIS 19 | 0.79 | | |
| MFIS 20 | 0.76 | 0.87 | 0.82 |
| MFIS 21 | 0.60 | | |
| **Total** | 0.63 | 0.75 | 0.81 |

***Table S14. Factor Reliability.*** *Chronbach's alpha for each factor*

| | FA 1. All Items | | FA 2. Controlling for Multicoleariy and Cross-factor loadings | | FA 3. Controlling for Multicolineariy and Poor KMO Scores | |
|---|---|---|---|---|---|---|
| **Factor** | **Fatigue** | **Depressed Mood** | **Fatigue** | **Depressed Mood** | **Fatigue** | **Depressed Mood** |
| Raw α | 0.98 | 0.97 | 0.96 | 0.93 | 0.97 | 0.96 |

***Table S15. Bartlett factor scores.*** *Factor scores assigned to each participant based on their survey scale responses for all factor analyses.*

| | FA 1. All Items | | FA 2. Controlling for Multicolineariy and Cross-factor loadings | | FA 3. Controlling for Multicolineariy and Poor KMO Scores | |
|---|---|---|---|---|---|---|
| **Participant** | **Fatigue** | **Depressed Mood** | **Fatigue** | **Depressed Mood** | **Fatigue** | **Depressed Mood** |
| AA_MDD020 | 0.83 | 1.06 | 1.12 | 1.17 | 0.98 | 1.15 |
| AB_MDD033 | 1.32 | 0.61 | 1.45 | 0.66 | 1.38 | 0.86 |
| AR_MDD019 | 1.70 | 1.27 | 1.73 | 1.39 | 1.63 | 1.29 |
| AR_MDD034 | 1.04 | -0.03 | 1.12 | -0.06 | 1.11 | -0.06 |
| AW_MDD029 | 1.43 | 0.85 | 1.35 | 1.32 | 1.42 | 0.97 |
| BH_MDD036 | 0.92 | 0.71 | 0.80 | 0.79 | 0.78 | 0.97 |
| CC_MDD031 | -0.89 | 0.04 | -0.91 | -0.11 | -0.92 | 0.10 |
| EF_MDD032 | 1.94 | 1.55 | 1.94 | 1.23 | 1.89 | 1.66 |
| JA_MDD014 | -0.21 | -0.16 | -0.01 | -0.44 | 0.01 | -0.21 |
| KK_MDD026 | 0.07 | 0.01 | 0.10 | -0.26 | 0.33 | -0.04 |
| KM_MDD035 | 0.22 | 2.92 | -0.11 | 3.20 | -0.01 | 2.72 |
| LM_MDD013 | 0.94 | 0.61 | 0.97 | 0.24 | 1.03 | 0.61 |
| MP_MDD022 | 0.06 | -0.09 | 0.30 | -0.20 | 0.31 | -0.15 |
| MU_MDD025 | 1.91 | 2.72 | 1.86 | 2.70 | 2.08 | 2.48 |
| RL_MDD021 | 0.07 | 1.22 | 0.34 | 1.45 | 0.26 | 1.32 |
| RW_MDD027 | 1.98 | 2.22 | 2.03 | 2.02 | 1.87 | 2.36 |
| SB_MDD030 | 1.80 | 1.87 | 1.84 | 1.69 | 1.89 | 1.63 |
| SS_MDD023 | 0.39 | 0.95 | 0.41 | 1.06 | 0.32 | 1.14 |
| TS_MDD016 | 0.80 | 0.76 | 0.63 | 0.49 | 0.66 | 0.39 |
| TW_MDD018 | 0.62 | 0.59 | 0.37 | 0.63 | 0.37 | 0.69 |
| YZ_MDD015 | 1.17 | 0.75 | 1.13 | 0.79 | 1.18 | 0.92 |
| AP_CTL009 | -0.75 | -0.66 | -0.75 | -0.80 | -0.72 | -0.73 |
| AW_CTL024 | -0.02 | -0.71 | -0.10 | -0.66 | -0.06 | -0.68 |
| DF_CTL027 | 0.68 | -1.04 | 0.58 | -0.99 | 0.56 | -0.94 |
| DP_CTL003 | -0.61 | -0.70 | -0.64 | -0.57 | -0.65 | -0.61 |
| FK_CTL016 | -0.47 | -0.50 | -0.37 | -0.56 | -0.42 | -0.61 |
| FZ_CTL022 | -0.52 | -0.82 | -0.37 | -0.71 | -0.39 | -0.79 |
| GL_CTL006 | -0.94 | -0.83 | -0.93 | -0.77 | -0.95 | -0.83 |
| HM_CTL033 | -0.71 | -0.75 | -0.69 | -0.84 | -0.67 | -0.81 |
| HT_CTL021 | 0.13 | -0.55 | -0.03 | -0.61 | 0.00 | -0.61 |
| JJ_CTL019 | -0.70 | -0.78 | -0.75 | -0.65 | -0.78 | -0.77 |
| KM_CTL013 | -1.33 | -0.82 | -1.33 | -0.82 | -1.32 | -0.86 |
| MA_CTL008 | -0.20 | -0.81 | -0.30 | -0.80 | -0.26 | -0.81 |
| MD_CTL004 | -1.33 | -0.82 | -1.33 | -0.82 | -1.32 | -0.86 |

| | | | | | | |
|---|---|---|---|---|---|---|
| MN_CTL014 | -0.46 | -0.49 | -0.42 | -0.33 | -0.45 | -0.46 |
| MQ_CTL018 | -1.33 | -0.82 | -1.33 | -0.82 | -1.32 | -0.86 |
| MW_CTL001 | 0.65 | -0.29 | 0.53 | -0.49 | 0.56 | -0.33 |
| PN_CTL011 | -0.67 | -0.69 | -0.61 | -0.67 | -0.59 | -0.80 |
| RB_CTL012 | -1.21 | -0.51 | -1.28 | -0.34 | -1.33 | -0.42 |
| RK_CTL031 | -1.35 | -0.62 | -1.35 | -0.66 | -1.37 | -0.70 |
| SF_CTL007 | 0.29 | -0.66 | 0.36 | -0.51 | 0.33 | -0.62 |
| SG_CTL023 | 0.35 | -0.59 | 0.37 | -0.53 | 0.34 | -0.43 |
| SG_CTL028 | -0.93 | -0.89 | -0.93 | -0.85 | -0.94 | -0.89 |
| SH_CTL015 | -1.05 | -0.72 | -1.15 | -0.67 | -1.13 | -0.73 |
| SP_CTL010 | -0.27 | -0.80 | -0.39 | -0.77 | -0.42 | -0.85 |
| TB_CTL005 | -0.93 | -0.83 | -0.84 | -0.85 | -0.85 | -0.90 |
| VR_CTL017 | -1.27 | -0.36 | -1.29 | -0.50 | -1.26 | -0.52 |
| XC_CTL032 | -1.33 | -0.82 | -1.33 | -0.82 | -1.32 | -0.86 |
| YL_CTL026 | -1.29 | -0.77 | -1.33 | -0.82 | -1.27 | -0.79 |
| YT_CTL030 | -0.56 | -0.77 | -0.51 | -0.59 | -0.59 | -0.72 |

*Table S16. Model comparison of choices for those with MDD from different factor analyses*

| | **FA 1. All Items** | | **FA 2. Colinearity and X-Factors** | | **FA 3. Colinearity and Poor KMO** | |
|---|---|---|---|---|---|---|
| **Model** | **AIC** | **BIC** | **AIC** | **BIC** | **AIC** | **BIC** |
| **Null (Choice ~ Effort$^2$ + Reward)** | 344.2 | 356.4 | 344.2 | 356.4 | 344.2 | 356.4 |
| **Null + Fatigue** | 344.7 | 361.0 | 344.4 | 360.6 | 344.4 | 360.6 |
| **Null + Fatigue + Reward*Fatigue** | 344.2 | 364.5 | 342.6 | 362.9 | 343.2 | 363.5 |
| **Null + Fatigue + Effort$^2$*Fatigue** | 343.4 | 363.7 | 340.4 | 360.7 | 341.6 | 361.9 |
| **Null + Fatigue + Effort$^2$*Fatigue + Reward*Fatigue** | 344.9 | 369.2 | 341.8 | 366.2 | 343.1 | 367.5 |
| **Null + Depression** | 346.2 | 362.4 | 346.2 | 362.4 | 346.2 | 362.4 |
| **Null + Depression + Reward* Depression** | 348.1 | 368.4 | 348.1 | 368.4 | 348.1 | 368.4 |
| **Null + Depression + Effort$^2$* Depression** | 347.9 | 368.2 | 347.3 | 367.6 | 347.5 | 367.8 |
| **Null + Depression + Effort$^2$* Depression + Reward* Depression** | 349.7 | 374.0 | 349.3 | 373.7 | 349.5 | 373.9 |

***Table S17***. *Median of bootstrapped parameter estimates and 95% confidence intervals for the best model using factor scores from each factor analysis*

| | FA 1 | FA 2 | FA 3 |
|---|---|---|---|
| **$E^2$** | -3.37 [-5.35, -1.78] | -2.78 [ -4.64, -1.27] | -2.02 [ -4.99, -1.43] |
| **R** | 5.20 [ 4.13, 6.56] | 5.22 [ 4.15, 6.57] | 5.21 [ 4.15, 6.55] |
| **FATIGUE** | -0.82 [-1.86, 0.17] | -0.90 [ -1.95, 0.07] | -0.93 [ -2.01, 0.10] |
| **$E^2$:FATIGUE** | -2.99 [-6.22, -0.37] | -4.14 [-7.19, -1.48] | -3.69 [ -7.12, -0.88] |
